# Evaluation of six different tests for *Schistosoma haematobium* diagnosis in a near-elimination setting: a prospective observational diagnostic accuracy study

**DOI:** 10.64898/2026.02.23.26346882

**Authors:** Naomi C. Ndum, Said M. Ali, Mohammed N. Ali, John Bergelin, Jan Hattendorf, Pytsje T. Hoekstra, Lisette van Lieshout, Tom Pennance, Khamis R. Suleiman, Jürg Utzinger, Peter Ward, Bonnie L. Webster, Stefanie Knopp

## Abstract

**Background:** Accurate diagnostic tools are needed in schistosomiasis elimination settings to determine prevalence thresholds for assigning or stopping interventions, guide pre- and post-elimination surveillance, and verify whether elimination has been reached. We assessed the accuracy of six different diagnostic tests in Pemba, Tanzania, a setting approaching *Schistosoma haematobium* elimination.

**Methodology:** A prospective diagnostic accuracy study was conducted from February to April 2025. From an initial cross-sectional single-day urine filtration (UF)-microscopy screening of 784 students, 69 *S. haematobium*-positive and 212 negative students were randomly selected for longitudinal sample collection. Four additional urine samples collected over four different days, were available from 262/281 participants and analysed by UF-microscopy. One sample per participant was analysed in parallel with five additional diagnostics: microscopy-based artificial intelligence (AI)-scanner, *Schistosoma*-ITS-2 qPCR, *S. haematobium*-Dra-1 recombinase polymerase amplification (RPA), Hemastix reagent strips, and up-converting particle lateral flow circulating anodic antigen assay (UCP-LF CAA). We assessed the sensitivity and specificity of the different diagnostics, using 5-day UF-microscopy examinations as reference test.

**Principal Findings:** A total of 85/262 participants were *S. haematobium*-positive using 5-day UF-microscopy. Directly compared with the reference test, the sensitivity for single-sample examination was: AI-scanner: 76.7% (95% confidence interval (CI): 71.0-82.5%); qPCR: 76.0% (95% CI: 70.1-81.4%); UF-microscopy: 61.2% (95% CI: 55.3-67.1%); RPA: 56.1% (95% CI: 50.0-62.2%); Hemastix: 44.6% (95% CI: 38.5-50.7%); and UCP-LF CAA: 30.6% (95% CI: 24.9-36.3%). Sensitivity increased with increasing infection intensity. The specificity of all investigated diagnostics was >92%, except for qPCR and RPA.

**Conclusion:** In near-elimination settings, multiple-day urine examination with standard UF-microscopy substantially improves case detection but is operationally challenging. For single-sample testing, among the six diagnostics investigated, the AI-scanner proved to be the most accurate. Hence, the AI-scanner might offer a promising alternative for research, clinical and programme use, but requires further validation in other settings and cost-effectiveness analyses.

**Trial registration:** clinicaltrials.gov, NCT06808750. Registered 08 January 2025, https://clinicaltrials.gov/study/NCT06808750.

**Author Summary:** As countries progress towards the schistosomiasis elimination goals set by the World Health Organization for 2030, case numbers and infection intensities decrease, which brings about diagnostic challenges. Indeed, accurate diagnostic tools are needed to precisely determine where to assign or stop interventions, implement test-and-treat approaches, and enable verification of elimination plus surveillance. We conducted the first prospective, head-to-head accuracy evaluation of six diagnostic tests, ranging from standard urine filtration (UF-) microscopy and haematuria assessment to advanced molecular and antigen tests and a new artificial intelligence (AI-) scanner for egg microscopy, all evaluated using the same urine sample, for *S. haematobium* detection in a near-elimination setting. We showed that examining multiple-day urine samples from the same individual with standard UF-microscopy, substantially increased the number of cases, especially when infection intensities were light. With 5-day UF-microscopy as the reference test, the specificity of all investigated single-sample tests was >92%, except for qPCR and *S. haematobium*-Dra-1 recombinase polymerase amplification. Sensitivity was >60% for the novel AI-scanner, qPCR and UF-microscopy. Sensitivity of all tests increased with increasing infection intensity. Schistosomiasis control and elimination programmes relying on single-sample UF-microscopy risk missing a substantial proportion of infections, impeding intervention decisions and jeopardising elimination targets. Multiple-day UF-microscopy improves case detection but is operationally challenging. For single-sample testing in near-elimination settings, an AI-scanner offers a promising alternative for research, clinical and programme use, warranting further cost-effectiveness and implementation studies.

## Introduction

Schistosomiasis is a neglected tropical disease (NTD) caused by infection with trematodes of the genus *Schistosoma*. The pathophysiology is mostly related to the host’s immune reaction to the innumerable eggs produced by female worms [1]. The global burden of schistosomiasis was estimated at 1.86 million disability adjusted life years lost in 2021 [2].

The World Health Organization (WHO) 2021-2030 Roadmap for NTDs sets the goal to eliminate schistosomiasis as a public health problem (<1% heavy intensity infections) globally and to interrupt transmission (absence of infections in humans) in selected areas by 2030 [3]. To achieve these goals in areas with an infection prevalence <10%, WHO suggests continuing population preventive chemotherapy with praziquantel at the same or reduced frequency as previously implemented in existing control programmes, or to use a clinical approach of test-and-treat [4,5]. In areas that have achieved interruption of transmission, elimination needs to be verified, and post-elimination surveillance implemented [4].

To determine prevalence thresholds to assign or stop interventions, implement test-and-treat approaches, and enable the verification of elimination and pre- and post-elimination surveillance, accurate, applicable, and reliable diagnostic tools are needed [1,6,7]. Standard diagnostics used in urogenital schistosomiasis control programmes, such as single urine filtration (UF)-microscopy and haematuria reagent strips, have a low sensitivity for light intensity infections (<50 *Schistosoma haematobium* eggs/10 mL of urine), which are associated with elimination settings [7-9]. Advanced diagnostics such as antigen and molecular tests and new artificial intelligence (AI)-based egg microscopy hold promise for a higher sensitivity and/or specificity, but prospective accuracy studies evaluating several of these diagnostics in parallel have not been conducted in near-elimination settings.

This study assessed accuracy of six diagnostic tests, ranging from standard UF-microscopy and haematuria assessment to advanced molecular and antigen tests to a new AI-scanner for egg microscopy, in a setting approaching *S. haematobium* elimination.

## Methods

### Ethics

The study protocol was approved by the Ethics Committee of Northern and Central Switzerland (AO_2024-00105) and the Zanzibar Health Research Institute (ZAHREC/01/PR/JAN/2025/01). It was prospectively registered at Clinical Trials (https://clinicaltrials.gov/study/NCT06808750). All study participants submitted a written informed consent signed by their legal guardian/parent and students aged 12-17 years an additional assent form signed by themselves. All children who tested *S. haematobium*-positive were informed about their result and treated with praziquantel (40 mg/kg) at the end of the study.

### Study design

This was an observational study, which consisted of (i) an initial screening for *S. haematobium* infections in two schools in Pemba Island, Tanzania, following a cross-sectional design to identify the required sample size of 60 positive and 190 negative individuals for the diagnostic study; (ii) a longitudinal collection of four additional urine samples across different days from all selected *S. haematobium*-positive and negative students to use combined results of the five UF-microscopy examinations per participant as reference test; and (iii) a prospective diagnostic accuracy study to assess the specificity and sensitivity of the investigated tests for the diagnosis of *S. haematobium* infections in low-prevalence settings approaching elimination. The study was conducted at the Public Health Laboratory-Ivo de Carneri (PHL-IdC) in Pemba. Pemba has achieved elimination as public health problem in many areas and is committed to achieve interruption of transmission [10,11]. Participants were recruited in two schools from February till March 2025. Laboratory examinations were conducted from February till April 2025.

### Determination of sample size

To estimate the required sample size to determine sensitivity and specificity for a diagnostic test using a gold standard reference test, we used the normal approximation for single proportions. Assuming a true specificity and sensitivity of the comparative diagnostic tests of 95%, we required approximately 70 *S. haematobium* egg-positive urine samples to estimate the sensitivity with a precision, defined as half the length of the 95% confidence interval, of approximately 5 percentage points. A total of 180 egg-negative participants are sufficient to estimate the specificity with a precision of 3.3 percentage points.

Based on previous studies in Pemba, we assumed that our two target study schools had a *S. haematobium* prevalence of ∼5% by single UF-microscopy. Hence, to identify 60 *S. haematobium*-positive students in the initial cross-sectional screening (results after examination of one urine sample per participant with UF-microscopy) and subsequently the required sample size of 70 *S. haematobium*-positive participants after examination of five urine samples per participant with UF-microscopy, we estimated that we needed to screen 1200 students from two schools (600 students per school).

### Selection of participants

Students from two schools in Pemba were eligible to participate in the cross-sectional screening if they attended grade 3, 4, 5, or 6, submitted a written informed consent signed by their parent or legal guardian and an additional written assent signed by the participant if aged 12-17 years old. In School 1, each grade included four classes (A, B, C, and D). For randomization, a computer-generated list indicated the class (A, B, C, or D) to include in the screening. All students present in the class at the day of registration were included. In School 2, there was only one class per grade and all students fulfilling the above criteria and present at the day of registration were included in the screening.

For longitudinal sampling and in the diagnostic accuracy study in School 1, all students that were *S. haematobium*-positive in the initial cross-sectional screening were included. Additionally, initially *S. haematobium*-negative students that were not randomly de-selected, were included (Fig 1; S1 Fig). In School 2, which had a higher *S. haematobium* prevalence than expected, *S. haematobium*-positive and negative students were included, if they were not randomly de-selected.

**Fig 1.**
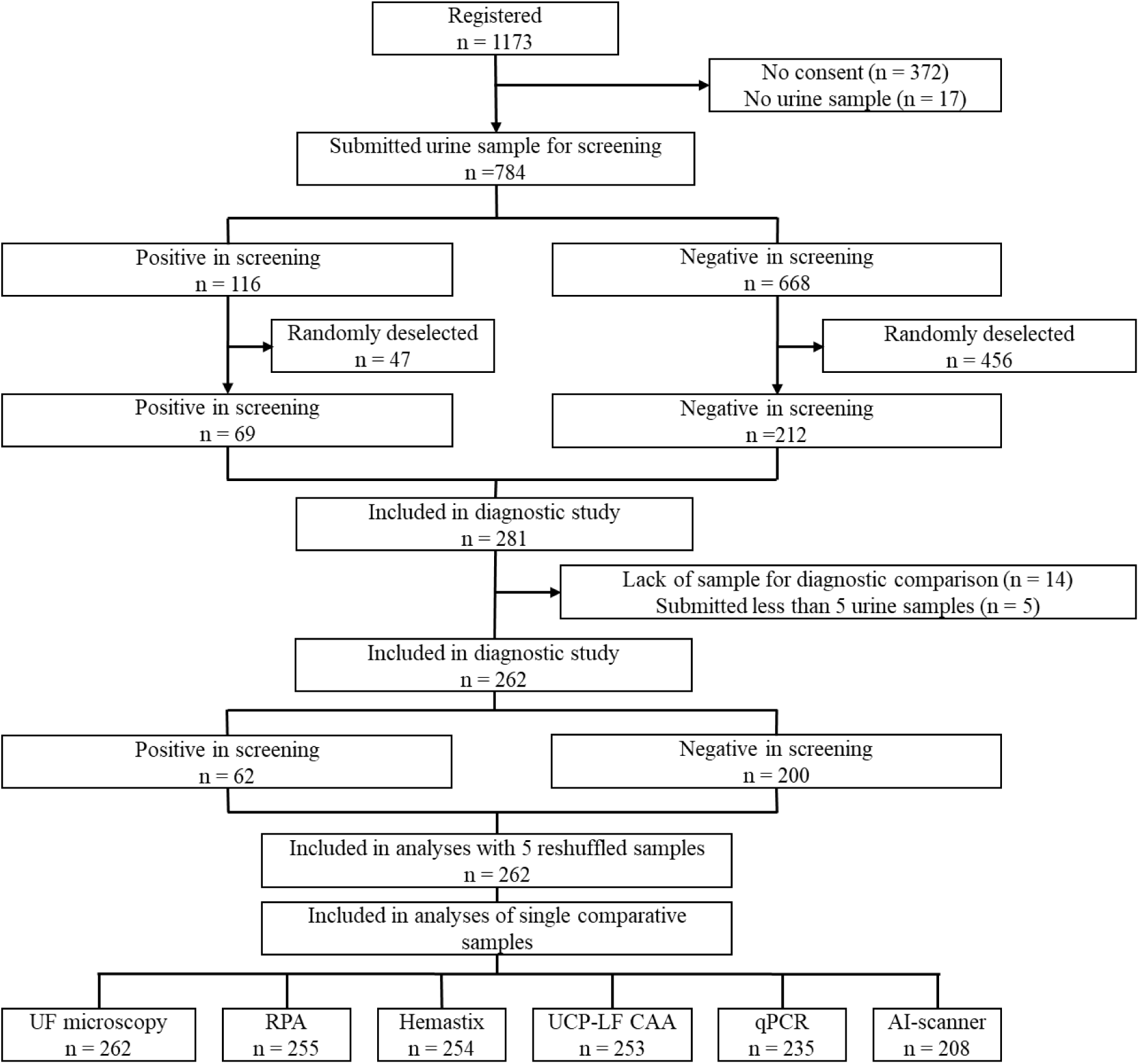
Inclusion and exclusion of participants in a prospective diagnostic accuracy study for *S. haematobium* in Pemba, Tanzania, in 2025. AI-scanner: artificial intelligence-based slide scanner; Hemastix: reagent strips; qPCR: *Schistosoma* ITS-2 real-time PCR; RPA: *S. haematobium* Dra1 recombinase polymerase amplification; UCP-LF CAA: up-converting reporter particle lateral flow circulating anodic antigen assay; UF-microscopy: urine filtration microscopy. More details are provided in S1 Fig.

### Demographic data and sample collection

Demographic characteristics of students eligible for the initial screening were recorded with Open Data Kit software (ODK; www.opendatakit.org) installed on tablet computers (Samsung Galaxy Tab A 2019). Gender was recorded by the interviewers, based on the students’ name and school uniform. On each day of sample collection, all students who met the inclusion criteria were handed a clean plastic container (100 mL) with a screwable lid and labelled with a personal identifier code and asked by the field enumerators to fill the container with their own urine sample. All urine samples were transported to the PHL–IdC, where they were examined with the following diagnostic procedures.

### Diagnostic procedures

Urine samples for the initial screening and longitudinally for an additional four days were collected from the study participants between 10 a.m. and 2 p.m. All urines samples were examined for *S. haematobium* infection using standard UF-microscopy based on filtration of 10 mL of mixed urine [12]. Among the longitudinally collected urine samples, one urine sample per participant was further tested with five additional diagnostics. Diagnostic procedures are described in detail below. Sample processing procedures for all diagnostic tests were started on the day of sample collection.

### Urine filtration (UF)-microscopy

UF-microscopy for *S. haematobium* egg detection in urine: UF-microscopy was performed at the PHL-IdC with all fresh urine samples of sufficient volume. Before it was subjected to filtration, the urine sample was properly mixed using a clean 10 ml plastic syringe. Subsequently, 10 mL of urine were drawn into the syringe and filtered through a 13 mm diameter filter (Sefar Nitex 03-25/19; Sefar, Lancashire, UK) that was held in a clean 13 mm diameter Swinnex filter holder (Millipore, Watford, UK). The filter disk was placed egg side up on a microscope slide and covered with glycerol-soaked cellophane. Subsequently, 1-3 drops of Lugol’s iodine were placed on the hydrophilic cellophane to stain the internal structures of any *S. haematobium* eggs making them more visible. The glass slides were placed under a light microscope at 20x magnification and the filter examined for *S. haematobium* eggs. If *S. haematobium* eggs were detected, their number was counted and recorded. If no *S. haematobium* eggs were detected, the value “0” was recorded for the respective sample.

### Artificial intelligence (AI)-scanner

AI-scanner for *S. haematobium* egg detection in urine: The Enaiblers AI slide scanner (Raven; EA-PROTO-01-P3.3) is a purpose-built whole-slide imager for automated reading of standard microscope glass slides in NTD programmes [13]. Originally developed to meet WHO diagnostic Target Product Profile requirements for soil-transmitted helminths (STH) [14], it has undergone extensive field validation for STH [15,16]. The study presented here is the first to assess its performance for *S. haematobium* egg detection. Slides were prepared as for standard UF-microscopy and imaged across the entire filter area using the AI-scanner at PHL-IdC. The scanner’s automated stage captured depth-stacked tile images to visualise eggs located below, within, and above the filter mesh. The onboard AI model (pipeline version 20250217t0725z_yolo8_detection_schistosoma_urine) processed the images in real time to identify candidate *S. haematobium* eggs (bounding boxes with an internal confidence score). Model parameters and the decision threshold were frozen before analysis of the first sample and not adjusted thereafter. In a human-in-the-loop verification step, all AI-detected candidates, ordered by model confidence, were reviewed in Oden software (version 10.2) by a trained operator, who classified each candidate as *S. haematobium* egg or non-egg material (e.g. debris, air bubbles, artefacts). Verified egg counts were then generated and reported per slide. Thus, the reported performance reflects the combined system (AI algorithm plus human verification), not a fully autonomous AI model. No generative AI or prompt-based inputs were used; the AI model was a pre-trained object detection algorithm optimised for parasitological image analysis. A sample was considered positive, if at least one *S. haematobium* egg was identified in the filter area.

### Real-time PCR (qPCR)

qPCR for amplification and detection of the *Schistosoma*-specific ITS-2 (Internal Transcribed Spacer) target (Sch-ITS-qPCR): Spin column-based DNA extraction from urine sediments was performed according to previous publications with some minor adaptations [17,18]. In brief, within two hours of receiving the fresh urine samples at the PHL-IdC and following thorough mixing, a 10 mL aliquot was taken and centrifuged in 15 ml Falcon tubes for five minutes at ≈700 x g to form a pellet. Half of the 1 mL pellet was frozen for at least 60 minutes at -80°C, while the other half was stored as a back-up sample. Following thawing, an 0.20 mL aliquot of the pellet was mixed with 0.8 gram of 1.4 mm Ceramic beads (Qiagen, Hilden, Germany). The sample was then bead-beaten (2x 5 minutes) using the “Qiagen TissueLyser LT” at a speed of 50 oscillations/second. Following a second freezing step (60 minutes at -80°C or overnight at -20°C), each sample was heated at 100°C for 10 minutes, followed by treatment with proteinase K for 2 hours at 55°C. Thereafter DNA extraction was performed using the QIAamp DNA mini kit (Qiagen, Hilden, Germany) per manufacturer’s instructions, with a final DNA elution of 200 µl. As part of the DNA extraction procedure a fixed volume of Phocid Herpesvirus-1 – PhHV (European Virus Archive Global – EVAg, cat. 011V-00884; Erasmus MC Rotterdam) was added to the isolation lysis buffer to act as internal qPCR control and for the detection of potential inhibition of amplification.

The extracted DNA was stored at 4°C and shipped, within 10 days, from PHL-IdC to Leiden University Medical Centre (LUMC) for Sch-ITS-qPCR analysis as previously described [17,19]. A single Sch-ITS-qPCR was performed on each DNA extraction using a CFX real-time detection system (Bio-Rad Laboratories) with 50 PCR amplification cycles run per sample. Results were reported as cycle threshold (Ct) values, meaning the amplification cycle at which the fluorescent signal exceeded the background level [17,19]. Negative and positive DNA controls were included in each set of samples run.

Since the implementation of the international Helminths External Molecular Assessment Scheme (HEMQAS), provided by the Dutch Foundation for Quality Assessment in Medical Laboratories (SKML), the LUMC-team have scored 100% in sensitivity and specificity of their *Schistosoma* qPCR in annual tests [20].

### Hemastix reagent strips

Hemastix reagent strips for microhaematuria detection in urine, as proxy for *S. haematobium* infection: Hemastix reagent strips (Siemens Healthcare, Zürich, Switzerland) were used for microhaematuria assessment in fresh urine samples at PHL-IdC. According to the manufacturer’s instruction, the reagent strip was dipped in the well-mixed urine sample and removed immediately. After 60 seconds, within 1-2 minutes after urine contact, the test pad was matched with the colour chart indicated on the manufacturer’s bottle and the result read and recorded. In line with the colour, the result was judged as either negative, trace, light (+), moderate (++), or high (+++). Samples were considered as *S. haematobium*-negative when microhaematuria was graded negative or trace, and as *S. haematobium*-positive when microhaematuria was light (+), moderate (++), or high (+++) [21].

### Recombinase Polymerase Amplification (RPA)

RPA for *S. haematobium* DNA detection from eggs in urine. RPA is an isothermal DNA amplification methodology to detect *S. haematobium* DNA from eggs within the fresh urine sample [22]. Ten mL of mixed urine were filtered as for UF-microscopy at PHL-IdC. The filter was then placed in a clean 1.5 mL Eppendorf tube. A crude DNA extraction procedure was performed on each sample using the SwiftDNA extraction kit (Xpedite Diagnostics, https://www.xpedite-dx.com/), with modifications to the recommended protocol to make it low-resource applicable. A volume of 100 µL of Buffer DL was added to the sample, along with 30 µL of SwiftDNA magnetic beads (Beads A). The mixture was vortexed for 30 seconds, briefly centrifuged, and then incubated at room temperature for at least five minutes. The mixture was then vortexed vigorously, centrifuged briefly and the Eppendorf tube was placed on a magnetic rack for at least one minute. The magnetic beads that capture sample impurities are attracted to the magnet, and the DNA elution (∼130 µL) was removed from the Eppendorf tube using a micropipette and stored in a clean 0.5 ml Eppendorf tube. All DNA elutions were stored at 4°C before being analysed. For every 25 samples extracted, a random negative (water) extraction control was included to enable the detection of DNA contamination during the extraction processes. All samples were analysed for the presence of *S. haematobium* DNA at PHL-IdC using the Sh-Dra1-RPA assay using Recombinase Aided Amplification (RAA) kits, which are used for the RPA reactions (Xpedite Diagnostics, <u>Halbergmoos, Germany</u>) following the manufacturer’s protocol. The Sh-Dra1-RPA assay was performed using 13.2 μL of each sample, in sets of 6 or 14 samples at a time, as described by Archer and colleagues [22]. A positive (*S. haematobium* extracted DNA) and negative (water) control was run within each set of samples run. Reactions were run on either an Axxin T8 (six samples and two controls) or T16 fluorometer (14 samples and two controls; Axxin, Eaglemont, Australia), respectively. Samples were automatically assigned by the fluorometer as positive or negative, based on pre-specified >200 Relative Fluorescence Units (RFU) rise in fluorescence above baseline.

### Up-converting reporter particle-lateral flow circulating anodic antigen assay (UCP-LF-CAA)

UCP-LF-CAA to detect *Schistosoma* antigen in frozen urine: The UCP-LF CAA assay was conducted at the PHL-IdC following the UCAA*hT*417 dry format as described previously [23]. Each run included a serial dilution of TCA-treated crude adult worm antigen (AWA-TCA) prepared in CAA-negative urine to validate the cut-off (2 pg/ml) and to quantify the CAA concentration in unknown samples. Samples with a CAA concentration above the pre-specified cut-off of 2 pg/ml were classified as positive [23].

### Blinding

All laboratory tests were conducted in separate laboratory rooms at PHL–IdC and the experts and assistants conducting the tests were blinded to the results of all tests and associated data.

### Objectives

The primary objective was to assess the specificity and sensitivity of all investigated diagnostic tests, using the combined results of standard UF-microscopy conducted on five urine samples collected over five different days as reference test. Further objectives included: to assess (i) specificity and sensitivity of all investigated tests using the UF-microscopy results of the same single sample as reference; (ii) specificity and sensitivity of all investigated tests with latent class analyses (LCA); (iii) change in proportion of *S. haematobium* infections when 5-day urine samples per individual were analysed with UF-microscopy; and (iv) sensitivity of all investigated diagnostic tests in relation to *S. haematobium* egg counts derived from UF-microscopy of the same sample or mean egg counts from 5-day samples, respectively.

### Statistical analysis

If a participant did not submit the urine sample for the different diagnostic tests, they were excluded from statistical analyses. Moreover, if a participant submitted fewer than five urine samples, they were excluded from statistical analyses.

If a participant submitted more than five urine samples, only the sample that was subjected to multiple diagnostic tests plus four additional samples that were randomly selected and reshuffled were included in statistical analyses. These five samples could include or not include the sample analysed in the initial screening. Of note, six or seven samples were collected and examined by UF-microscopy from several children due to a “too” rigorous mop-up of children with missing samples.

Samples selected as described above and examined with UF-microscopy and one or more of the diagnostic tests under evaluation were included in statistical analyses.

Participants’ sociodemographic and baseline characteristics were analysed descriptively. First, the primary outcomes (i.e. the specificity and sensitivity of all investigated diagnostic tests) were calculated using 5-day UF-microscopy results as the reference test for direct comparison. Second, the specificity and sensitivity of all investigated diagnostic tests were calculated using the UF-microscopy results from the same single sample as reference test for direct comparison. Third, specificity and sensitivity were estimated using a Bayesian LCA model for two dependent tests in two populations without a gold standard [24]. Since this model is overparameterized, it requires the specification of at least two informative priors for identifiability. False-positive results are very rare for UF-microscopy. Hence, we specified informative priors for specificity and sensitivity. As specificity of UF-microscopy is close to perfect, we assumed with 95% certainty that the specificity is at least 99.5%. In addition, the informative prior for the sensitivity of UF-microscopy was determined as follows: First, we estimated the apparent proportions of positives from one up to five UF-microscopy results. To do so, we repeatedly and randomly selected the required number of samples from among all students who provided at least five urine samples. Next, we fitted an Emax model to the observed proportions of positive samples to predict the proportion of positive samples that would have been obtained if 20 urine samples per person had been assessed. This value and its corresponding 95% confidence interval (CI) were converted into UF-microscopy sensitivity. The CI of the sensitivity was used as prior for the Bayesian model. Six separate Bayesian models were run, each time using one of the investigated single-sample tests as first diagnostic approach and five UF-microscopy results as the second diagnostic approach.

For additional secondary outcomes, the proportion of *S. haematobium* infections and intensity of infections based on UF-microscopy was calculated cumulatively across five samples. To calculate the cumulative increase in infection intensity across five samples and days, the intensity of the first positive sample per participant was kept across all subsequent days. The intensity of additional positive cases on the following day was added on the day of occurrence and kept across all subsequent days. The threshold for light intensity infections in each sample was 1-49 eggs/10 mL urine and for heavy intensity infection was ≥50 eggs/10 mL urine [4].

Moreover, the sensitivity of each investigated diagnostic test was determined in relation to *S. haematobium* egg counts, calculated using (i) the mean egg count of 5-day UF-microscopy results and (ii) the egg counts of the single sample that was subjected in parallel to the other diagnostic tests, using generalized linear models.

## Results

### Participant characteristics

A total of 1173 students from two schools were recruited for the cross-sectional study (Fig 1; S1 Fig). Among them, 784 students submitted a urine sample for *S. haematobium* screening using UF-microscopy. Among them, 281 students (69 *S. haematobium*-positive and 212 negative), were included in the longitudinal sampling and diagnostic accuracy study. Fourteen students did not provide a sample for the diagnostic study, and five students provided less than four additional samples; hence, they were excluded from further statistical analyses. A total of 262 students who submitted at least five samples were included in the analyses of the longitudinal sampling and diagnostic study, respectively. Due to insufficient urine sample volumes to run all tests in parallel on the same urine sample, most of the investigated tests had a smaller sample number than 262 (Fig 1).

Baseline characteristics of the 262 students selected for inclusion in the diagnostic study, included a median age of 11 years, and 138 (51.1%) males (Table 1). Among them, 62/262 were *S. haematobium*-positive in the screening and 51/262 had a light intensity infection. Mean and median egg counts of positives were 28.3 and 11 eggs/10 mL, respectively.

**Table 1.** Baseline characteristics of 262 participants selected for inclusion in a prospective diagnostic accuracy study conducted for *S haematobium* in Pemba, Tanzania, in 2025. IQR: interquartile range; UF: urine filtration.

|  |  | **Total**<br>(*n* = 262) |  |
| --- | --- | --- | --- |
| **Variable** |  | *N* | % |
| **Sex** | Female | 130 | 49.6 |
|  | Male | 132 | 50.4 |
| **Age (years)** | Median (LIQR–UIQR) | 11 (10-12) |  |
| ***S. haematobium* infection diagnosed with a single UF-microscopy in screening** | Positive in screening | 62 | 23.7 |
|  | Light intensity (1-49 eggs/10 mL urine) in screening | 51 | 19.5 |
| | Heavy intensity ($\geq 50$ eggs/10 mL urine) in screening | 11 | 4.2 |
|  | Mean egg count of positive (range) | 28.3 (1-253) |  |
|  | Median egg count of positive (IQR) | 11 (4-32) |  |

### Sensitivity and specificity of diagnostic tests

The accuracy indicators of all investigated tests, using 5-day UF-microscopy as the reference test, are presented in Table 2. Most tests had a specificity >90%, but qPCR and RPA showed lower values. Among all single-sample tests, the AI-scanner, qPCR and UF-microscopy had a sensitivity >60%. When the single UF-microscopy result of the same sample was used as reference test, only Hemastix and UCP-LF CAA had a specificity >90% (Table 2). All tests except the UCP-LF CAA had a sensitivity >60%. The LCA estimated a specificity >90% for all single-sample tests, except qPCR and RPA (Table 3). A sensitivity of >60% was estimated for the AI-scanner and qPCR, only.

**Table 2.** Accuracy of six diagnostic tests assessed in a prospective diagnostic accuracy study conducted for *S haematobium* in Pemba, Tanzania, in 2025. Diagnostic single-sample (1x) test results, sensitivity and specificity are presented when either the results of 5-day UF-microscopy examinations (5x), or from a single UF examination (1x) from the same single sample are used as reference test.

|  |  | 5x UF-microscopy |  |  | 1x UF-microscopy<br>(same sample) |  |  |
| --- | --- | --- | --- | --- | --- | --- | --- |
|  |  | Negative | Positive | Total | Negative | Positive | Total |
| **1x AI-scanner** | **Negative** | 125 | 17 | 142 | 139 | 3 | 142 |
|  | **Positive** | 10 | 56 | 66 | 22 | 44 | 66 |
|  | **Total** | 135 | 73 | 208 | 161 | 47 | 208 |
|  | **Sensitivity (95% CI)** | 76.7% (71.0-82.5%) |  |  | 93.6% (90.3-96.9%) |  |  |
|  | **Specificity (95% CI)** | 92.6% (89.0-96.2%) |  |  | 86.3% (81.7-91.0%) |  |  |
| **1x qPCR** | **Negative** | 121 | 19 | 140 | 138 | 2 | 140 |
|  | **Positive** | 35 | 60 | 95 | 48 | 47 | 95 |
|  | **Total** | 156 | 79 | 235 | 186 | 49 | 235 |
|  | **Sensitivity (95% CI)** | 76.0% (70.5-81.4%) |  |  | 95.9% (93.4-98.5%) |  |  |
|  | **Specificity (95% CI)** | 77.6% (72.2-82.9%) |  |  | 74.2% (68.6-79.8%) |  |  |
| **1x UF-microscopy** | **Negative** | 177 | 33 | 210 |  |  |  |
|  | **Positive** | 0 | 52 | 52 |  |  |  |
|  | **Total** | 177 | 85 | 262 |  |  |  |
|  | **Sensitivity (95% CI)** | 61.2% (55.2-67.1%) |  |  |  |  |  |
|  | **Specificity (95% CI)** | 100% (100-100%) |  |  |  |  |  |
| **1x RPA** | **Negative** | 140 | 36 | 176 | 159 | 17 | 176 |
|  | **Positive** | 33 | 46 | 79 | 45 | 34 | 79 |
|  | **Total** | 173 | 82 | 255 | 204 | 51 | 255 |
|  | **Sensitivity (95% CI)** | 56.1% (50.0-62.2%) |  |  | 66.7% (60.9-72.5%) |  |  |
|  | **Specificity (95% CI)** | 80.9% (76.1-85.8%) |  |  | 77.9% (72.9-83.0%) |  |  |
| **1x Hemastix** | **Negative** | 170 | 46 | 216 | 198 | 18 | 216 |
|  | **Positive** | 1 | 37 | 38 | 5 | 33 | 38 |
|  | **Total** | 171 | 83 | 254 | 203 | 51 | 254 |
|  | **Sensitivity (95% CI)** | 44.6% (38.5-50.7%) |  |  | 64.7% (58.8-70.6%) |  |  |
|  | **Specificity (95% CI)** | 99.4% (98.5-100.4%) |  |  | 97.5% (95.6-99.4%) |  |  |
| **1x UCP-LF CAA** | **Negative** | 162 | 59 | 221 | 190 | 31 | 221 |
|  | **Positive** | 6 | 26 | 32 | 11 | 21 | 32 |
|  | **Total** | 168 | 85 | 253 | 201 | 52 | 253 |
|  | **Sensitivity (95% CI)** | 30.6% (24.9-36.3%) |  |  | 40.4% (34.3-46.4%) |  |  |
|  | **Specificity (95% CI)** | 96.4% (94.1-98.7%) |  |  | 94.5% (91.7-97.3%) |  |  |

**Table 3.** Bayesian latent class analyses for six diagnostic single-sample tests (1x) for *S. haematobium* investigated in a prospective diagnostic study conducted in Pemba, Tanzania, in 2025.

|  | **Test sensitivity**<br>(95% CI) | **Test specificity**<br>(95% CI) |
| --- | --- | --- |
| **1x qPCR** | 75.8%<br>(64.4-84.6%) | 86.5%<br>(76.0-95.7%) |
| **1x AI-scanner** | 66.4%<br>(55.5-77.6%) | 93.8%<br>(87.4-98.8%) |
| **1x RPA** | 56.0%<br>(43.9-67.1%) | 85.1%<br>(76.3-92.7%) |
| **1x UF-microscopy** | 50.0%<br>(39.9-60.5%) | 98.7%<br>(95.2-99.9%) |
| **1x Hemastix** | 38.1%<br>(29.0-47.9%) | 99.1%<br>(96.6-100%) |
| **1x UCP-LF CAA** | 28.1%<br>(19.6-38.5%) | 96.7%<br>(92.4-99.6%) |

### Impact of multiple sampling on proportion of detected *S. haematobium* infections

Fig 2 shows that 5-day UF-microscopy increased the cumulative proportions of *S. haematobium* infections. While after reshuffling of samples the analyses of a single sample revealed that 19.9% (52/262) of individuals were infected, the examination of 5-day samples revealed that 32.4% (85/262) were infected. Light intensity infections increased from 13.0% (34/262) to 23.3% (61/262) and heavy intensity infections from 6.9% (18/262) to 9.2% (24/262), respectively.

**Fig 2.**
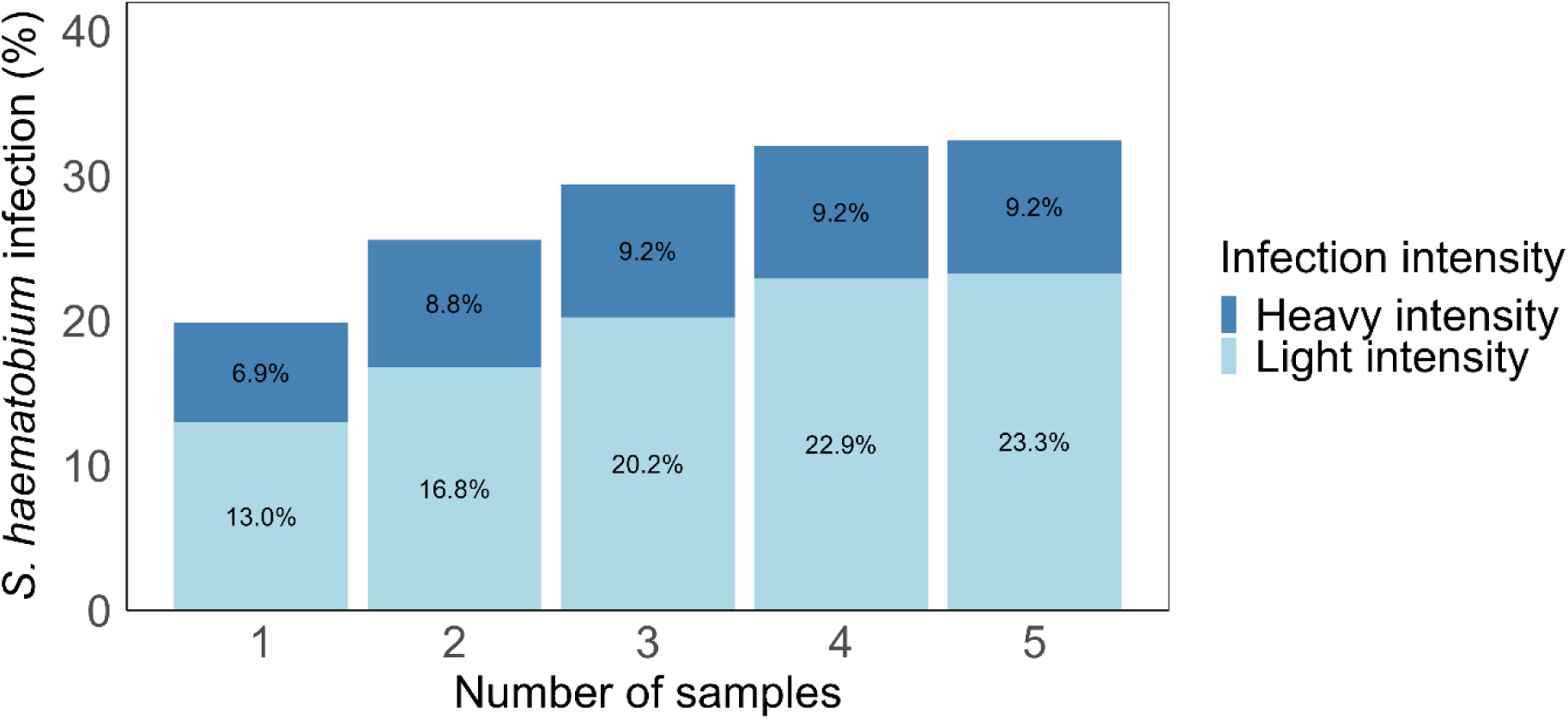
Cumulative proportion of *S. haematobium* infections and infection intensities in a prospective diagnostic accuracy study conducted in Pemba, Tanzania, in 2025. Cumulative proportion of *S. haematobium* infections and infection intensities when one to five samples were examined with urine filtration microscopy.

### Sensitivity of diagnostic methods in relation to *S. haematobium* egg counts

The sensitivity of all investigated diagnostics increased with increasing *S. haematobium* egg counts (Fig 3). In comparison with the results of 5-day UF-microscopy (Fig 3A), a sensitivity of at least 75% was achieved by qPCR and AI-scanner at counts of >5 eggs/10 mL urine, by single-sample UF-microscopy at counts of >20 eggs/10 mL urine, and for all other investigated tests at counts of >100 eggs/10 mL. In comparison with the results of the single UF-microscopy performed on the same sample as the other single-sample tests (Fig 3B), the sensitivities of the investigated tests showed similar increasing trends. The mean and median egg counts derived by 5-day and single-sample UF microscopy, respectively, are presented in Table S1.

**Fig 3:**
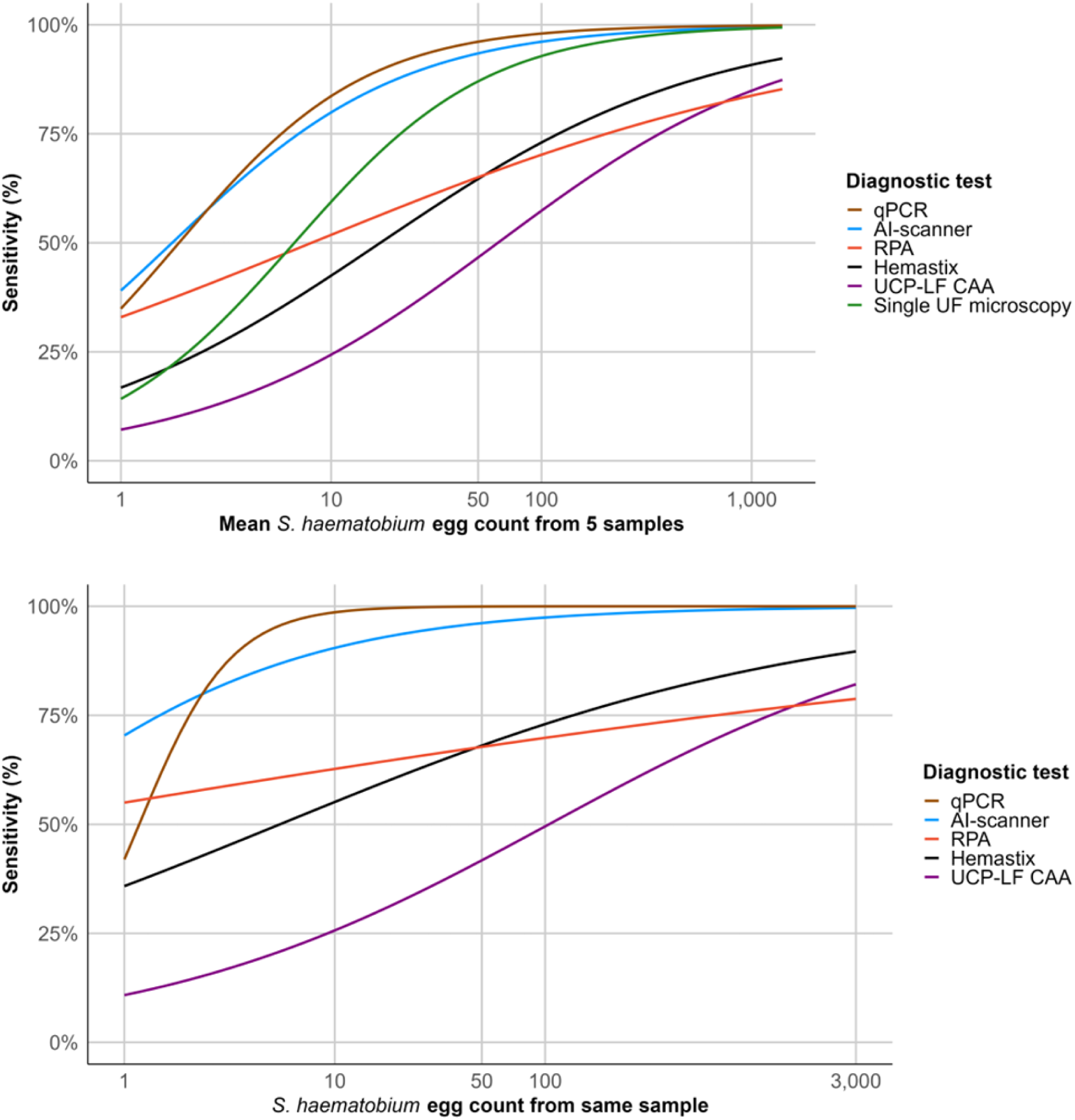
Sensitivity of single-sample diagnostic tests for *S. haematobium* according to infection intensity (egg counts) in a prospective diagnostic accuracy study conducted in Pemba, Tanzania, in 2025. (A) Sensitivity of each test when compared with the 5-day urine filtration (UF)-microscopy as reference test. All investigated diagnostic tests were performed on a single urine sample per participant; infection intensity on the x-axis represents the mean *S. haematobium* egg count per 10 mL urine across 5-day UF-microscopy examinations (reference test). (B) Sensitivity of each test when compared with single-sample UF-microscopy as reference test. All diagnostic tests and UF-microscopy were performed on the same urine sample; infection intensity on the x-axis represents the *S. haematobium* egg count per 10 mL urine from that same-sample UF-microscopy examination. Sensitivity (%) is shown on the y-axes and egg counts (eggs/10 mL urine; log10 scale) on the x-axes. AI-scanner: artificial intelligence-based slide scanner; Hemastix: reagent strips; qPCR: *Schistosoma* ITS-2 real-time PCR; RPA: *S. haematobium* Dra1 recombinase polymerase amplification; UCP-LF CAA: up-converting reporter particle lateral flow circulating anodic antigen assay; UF-microscopy: urine filtration microscopy.

## Discussion

As countries progress towards the schistosomiasis elimination goals set for 2030, prevalence and infection intensities decrease, which poses a challenge for accurate diagnosis [25]. Yet, accurate diagnostic tools are required in elimination settings to determine prevalence thresholds for intervention assignment, to verify elimination, and for pre- and post-elimination surveillance. We conducted the first prospective, head-to-head accuracy evaluation of six tests for *S. haematobium* diagnosis, all evaluated in parallel using the same urine sample, in a near-elimination setting.

Among the investigated tests, the AI-scanner, assessed here for the first time for *S. haematobium* diagnosis, showed promising results with a specificity of 92.6% and sensitivity of 76.7%, as revealed by direct comparison with 5-day UF-microscopy results as reference test. Both accuracy indicators are expected to increase by using larger and more diverse training datasets, with further enhanced image pre-processing to better distinguish eggs from background material. The specificity (77.6%) and sensitivity (76.0%) of qPCR are in line with those of previous studies [17,26,27]. The RPA had a similar specificity (80.9%) to the qPCR, but considerably lower sensitivity (56.1%). The difference in sensitivity likely reflects the different sample preparation and DNA extraction procedures used for the respective tests [28]. The qPCR used a high resource multiple-step column DNA extraction protocol with bead beating of the sediment of 10 mL of urine, while the RPA employed a crude DNA extraction method (simple lysis) from eggs trapped on a filter from 10 mL of urine. Clearly, the latter sample preparation method needs proper optimisation and evaluation [29]. The Hemastix reagent strips showed a low sensitivity (44.6%), but a surprisingly high specificity (99.4%) in the current study, which is in contrast to a previous study conducted in Zanzibar [21]. Of note, this method assesses haematuria as a proxy for *S. haematobium* infection. Since haematuria is related to the number of eggs excreted [30], and infection intensities were mostly very low in our near-elimination setting, the observed low sensitivity is not surprising. The high specificity may be explained by considering only samples with definite microhaematuria but not trace results as positive in our study and is in line with results from a recent systematic review [7]. The UCP-LF CAA had a high specificity (96.4%), but a relatively low sensitivity (30.6%). These results are in contrast to previous studies in high- but also low-endemic settings, where the UCP-LF CAA showed high values for both specificity and sensitivity [31-33]. Reasons for the observed low sensitivity may include the use of a lower volume of urine compared to a previous study conducted in the same area, in which the most sensitive format of the UCP-LF CAA test used 2 mL of urine [31], but also additional yet unidentified factors may have played a role. The accuracy indicators could also have been influenced by different study design and participant selection criteria applied in the current study compared with previous studies. Importantly, the diagnostic methods used here targeted different biomarkers of infection. While both UF-microscopy and AI-scanner use the same target, *S. haematobium* eggs, the UCP-LF CAA assay detects antigen excreted by worms, and the molecular methods (qPCR and RPA) detect DNA extracted from eggs, rendering direct comparisons between tests more challenging.

Notably, the estimates of our accuracy indicators derived by direct method comparison must be interpreted in light of the limited sensitivity of UF-microscopy, even when five samples were examined. For tests that are at least as sensitive as UF-microscopy, such as the AI-scanner and qPCR in our study, using UF-microscopy as the reference test inevitably classified some true infections as “false positive”. This effect was particularly visible in the comparison of the AI-scanner with UF-microscopy, where several samples were recognized as negative by UF-microscopy, despite the presence of operator-verified *S. haematobium* eggs on the digitised images of the AI-scanner. Consequently, the specificities reported for the AI-scanner, and likely also qPCR, can be regarded as conservative and partly reflect missed infections by the reference test rather than true biological false positives. To account for the imperfect sensitivity of UF-microscopy, in addition to the direct method comparison, we used a LCA model incorporating a prior for sensitivity reflecting the proportion of positives when 20 urine samples per person had been assessed with UF-microscopy. The LCA revealed a higher specificity (86.5%) and similar sensitivity (75.8%) for qPCR and a similar specificity (93.8%) and lower sensitivity (66.4%) for the AI-scanner, compared with direct method comparison.

In addition to the results pertaining to the accuracy indicators of the different diagnostics, we found that when urine samples from the same participants were collected and examined with standard UF-microscopy over multiple days, the proportion of *S. haematobium* infections detected increased considerably. The increase was most prominent for light intensity infections. Similar observations have been made elsewhere and for other parasites [30,34,35]. Important implications for schistosomiasis control and elimination need to be drawn. Settings that have (achieved) an apparently low prevalence of infection based on a single UF-microscopy may not be selected for preventive chemotherapy according to WHO guidelines [4]. Yet, the true prevalence is likely higher, and hence, there is a risk of (i) leaving a considerable number of infected individuals untreated and (ii) a rebound of infections due to an underestimated transmission force jeopardizing the elimination goals. Hence, to accurately estimate the “true” prevalence of infection on which intervention decisions are taken, multiple samples per individuals should be examined with UF-microscopy. However, a multiple sampling approach may be operationally difficult for monitoring and evaluation of programmes’ impact and particularly for (passive) surveillance conducted in health facilities in low-resource settings. Alternatively, highly accurate diagnostic tests should be applied for single-sample testing.

Among the tests that were investigated as alternative to UF-microscopy in this study, when assessing sensitivity in relation to *S. haematobium* egg counts, only the qPCR and AI-scanner were able to detect counts of >5eggs/10 mL urine with a sensitivity >75%. All other investigated tests were only able to detect higher intensity infections (often only with more than several hundred eggs/10 mL) with sufficient sensitivity. These findings suggest that for elimination settings, and depending on the purpose, only the qPCR and AI-scanner may be considered useful tools.

Concluding, in settings nearing *S. haematobium* elimination, repeated samples need to be analysed with standard UF-microscopy to obtain accurate prevalence estimates for intervention decisions. However, collecting and examining samples over multiple days is operationally challenging. For single-sample testing, the AI-scanner showed promising specificity and sensitivity for *S. haematobium*. Including well trained software that is also applicable for diagnosis of other infections (e.g. *Schistosoma mansoni* and STHx in stool samples) [36], it might become a ready-to-use tool that can be set up in laboratories that are able to prepare slides for standard UF-microscopy. Hence, the AI-scanner might offer an alternative for research, clinical and programme use in near-elimination settings, but warrants further scientific inquiry and cost-effectiveness analyses before wider use.

## Data Availability

All relevant data are within the manuscript and its supporting information.

## Acknowledgments

The authors express their gratitude to all students who participated in this study and thank the teachers for their collaboration. We sincerely thank the field and laboratory teams of the PHL-IdC in Pemba, Tanzania, for their support of data collection and sample processing and examination. We extend our appreciation to Maureen Blokland-Flink and Céline Harmanus from LUMC whose support facilitated the examination of samples in this study. The authors acknowledge the unwavering support of Xpedite Diagnostics https://www.xpedite-dx.com/ for their support with the RPA methodologies and reagent supply.

## Author Contributions

Naomi C. Ndum: Funding Acquisition, Investigation, Data Curation, Formal Analysis, Visualization, Writing – Original Draft Preparation

Said M. Ali: Methodology, Project Administration, Supervision, Resources Mohammed N. Ali: Investigation

John Bergelin: Investigation

Jan Hattendorf: Conceptualization, Formal Analysis, Writing – Review & Editing, Resources

Pytsje T. Hoekstra: Conceptualization, Methodology, Investigation, Writing – Review & Editing, Resources

Lisette van Lieshout: Funding Acquisition, Conceptualization, Methodology, Investigation, Writing – Review & Editing, Resources

Tom Pennance: Investigation, Writing – Review & Editing, Resources Khamis R. Suleiman: Investigation

Jürg Utzinger: Supervision, Writing – Review & Editing

Peter Ward: Conceptualization, Methodology, Investigation, Writing – Review & Editing, Resources

Bonnie L. Webster: Conceptualization, Methodology, Investigation, Writing – Review & Editing, Resources

Stefanie Knopp: Funding Acquisition, Conceptualization, Methodology, Investigation, Data Curation, Project Administration, Supervision, Writing – Original Draft Preparation, Writing – Review & Editing, Resources

## Supporting information

**S1 Table. Characteristics of samples identified as positive with urine filtration-microscopy in a prospective diagnostic accuracy study conducted in Pemba, Tanzania, in 2025.**

**(PDF)**

**S1 Fig. Detailed flowchart of participant and sample selection in a prospective diagnostic accuracy study conducted in Pemba, Tanzania, in 2025.**

**(PDF)**

**S1 STARD Checklist. The filled checklist is based on the STARD Statement-Checklist of items that should be included in reports of diagnostic accuracy studies, developed by the STARD Initiative,** https://stard-statement.org/.

**(PDF)**

**S1 Dataset and dictionary (XLSX)**

